# Promoting helmet usage in undergraduates with electric and non-electric bicycles and scooters: a single-site survey and intervention

**DOI:** 10.1101/2024.12.19.24319348

**Authors:** Alanna Dorsey, Gabrielle Li, Spencer Cha, Victor Ritter, Enora Le Flao, David B. Camarillo

**Author notes:** Author information Alanna Dorsey Gabrielle Li Spencer Cha Victor Ritter Enora Le Flao David B. Camarillo. **Corresponding author:** Enora Le Flao.

## Abstract

College students report alarmingly low helmet usage while riding bikes and other open-wheeled forms of transportation (OWTs), increasing risk of brain injury and skull fracture. Despite prior interventions, this problem persists, necessitating an updated examination of the current barriers to helmet usage, and how to effectively mitigate them. This study aimed to determine the effect of class year on helmet usage in American college students, and explore emerging trends with electric bikes, skateboards, and scooters. We created and distributed an online survey to undergraduate students at Stanford University in California (N = 400) who regularly used electric or non-electric bikes, skateboards, or scooters. This survey collected information on helmet usage, attitudes on bike safety, and demographics, and the findings were used to design a pilot intervention. Students who regularly wore helmets were recruited as peer agents (N=3) and were trained to ask friends and classmates to pledge to wear a helmet. We observed bike, scooter, and skateboard rides (N = 4885), and followed up with students who pledged to analyze their behavior change. We confirmed prior trends that most college students never or rarely wear a helmet, and the most common reasons for not wearing a helmet were “Helmets mess up my hair” (44%), “I don’t see others wearing helmets” (43%), and “It’s unlikely I will fall/crash” (34%). We also found that helmet usage was similarly low (48% of participants reported never wearing a helmet) across class years, but class years reported different reasons for not wearing a helmet. Lastly, there was no difference in helmet usage between electric and non-electric OWT users for any of the frequency categories (p=0.43 for daily, p=0.44 for a few times a week, p=0.77 for a few times a month, p=0.23 for rarely, p=0.15 for never), revealing that previous trends in helmet behavior also apply to American college students with electric OWTs. The pilot intervention resulted in a non-significant yet positive trend, and students who pledged demonstrated a behavior change (as measured by follow-up surveys about helmet usage) that persisted 3 weeks after the conclusion of the intervention. In addition to finding that past trends about helmet behavior in college students are still present, we discovered new trends on electric OWTs and class years in American college students. We also found that our peer agent program shows promise in modifying the behavior of college students.

## Introduction

Electric bikes, skateboards and scooters have become rapidly popular and may become one of the most common forms of transportation in cities and college campuses. Thus, the unique challenges they pose will only become more pressing. The fast speeds and lack of regulation of electric open-wheeled forms of transportation (OWTs) endanger riders, and the incidence of injury is increasing precipitously. Data from the National Electronic Injury Surveillance System (NEISS) of the United States shows that there were over 45,000 e-bicycle injuries and nearly 15,000 e-scooter injuries in 2018, and a third of these were head injuries.^1,2^ The NEISS data also revealed that between 2014 and 2018, there was a 365% increase in emergency department admissions for e-scooter injuries. Compared to non-electric bikes, e-scooter riders were more likely to be admitted to a major trauma center and be diagnosed with a severe brain injury.^3^ By mile, there are more injuries when riding an e-scooter (180 injuries per million miles) than driving a motor vehicle (1 injury per million miles), thus electric scooters and bikes pose a safety issue.^4^ The electric OWT industry has a projected growth in the upcoming years and may result in more injuries if the lack of regulation and effective intervention are not addressed.^5^

College students comprise a large portion of electric OWT users,^6^ and thus are especially vulnerable to electric OWT-related injuries. Many university students ride electric and non-electric bikes, skateboards, or scooters, and their lack of helmet usage puts them at risk of head and brain injury. In the United States, about 2-22% of university commuters bike to class or work,^7,8^ of which about 9% have experienced a crash within the past year,^9^ resulting in potential head, neck, or brain injuries. Such trauma can be particularly debilitating for students – a majority of college students with traumatic brain injury report academic challenges, fatigue, and memory problems as a result of their injury, making academic success and overall well-being more difficult to achieve.^10^ Traumatic brain injuries sustained as a result of bicycle crashes present a pressing public health issue for universities.

Helmets are an efficient solution to prevent traumatic brain injury, yet many universities struggle to get students to wear them. Despite the clear benefits of wearing helmets – helmeted cyclists are 72% less likely to sustain a traumatic brain injury compared to non-helmeted cyclists^11^ – many college students neglect to wear them. At universities across the United States, about 60-76% of undergraduates never or rarely wear a helmet while commuting.^8,12,13^ One survey found that teenagers prioritize safety less than appearance and peer acceptance.^14^ This undervaluing may be attributed to young adults’ overconfidence in their own cycling ability.^15,16^ Other studies pointed more directly to the roles of social influence: college students wear helmets more frequently if they grew up in a bicycle-friendly community or had a norm of family helmet use.^13^ Surveys throughout the US have suggested other reasons, such as inconvenience, not owning a helmet, and forgetting to wear one.^8,12,16^ Furthermore, researchers have found that electric OWT users in general report low helmet usage, increasing their vulnerability to traumatic brain injury and skull fractures. Studies conducted in multiple countries found that of emergency department patients with e-scooter-related injuries, between 0-7% of them were wearing a helmet.^5^ Some researchers have explored the intention to wear a helmet for e-bike riders in Chinese colleges and e-scooter riders in U.S. colleges, but the rates of helmet usage have not been studied in American students with different forms of electric OWTs, to our knowledge.^6,17^ There is a need for more information on different types of open-wheeled transport. There is also a knowledge gap regarding the impact of class year on helmet usage, which could inform the development of more appropriate interventions suited for each class year.

Helmets prevent injury, and university stakeholders must address the alarmingly low helmet usage to promote student health. Understanding the reasons underlying the lack of helmet usage will allow the design of effective and tailored interventions. A previously published intervention addressed the trend of low undergraduates helmet usage through the use of peer agents.^18^ Psychosocial approaches are most effective to address health issues with significant social pressure,^19^ so researchers trained student cyclists as “peer agents” to ask students to sign a helmet “pledge card,” for which they would be rewarded with a free helmet.^18^ This peer-to-peer approach addressed the social aspect of helmet usage by counteracting negative peer pressure. It also educated students about their personal crash risk, utilized incentives, and increased accessibility of helmets. As a result of this multi-faceted approach, the intervention nearly doubled helmet usage rates at the university. However, this peer agent program has yet to be replicated at another university, to our knowledge.

The primary aims of the survey were to explore helmet usage in different class years and in those riding electric OWTs. We hypothesized that helmet usage beliefs and behaviors vary by class year – for instance, that first-year students wear helmets at a different frequency, or have different reasons for not wearing helmets. We also tested the hypothesis that students with electric forms of transport wore a helmet more often. The secondary aim was to evaluate past helmet programs at the campus and identify new avenues to disseminate information to students. Based on these findings, we then designed, implemented and evaluated a data-driven intervention. We hypothesized that implementing a 5-week pilot intervention consisting of social marketing, incentives, and education would increase helmet use among undergraduates, and that this change would be sustained for weeks after the intervention.

## Methods

### Helmet Behavior Survey

#### Study Design

A cross-sectional survey (titled “Caring for Your Brain: A Survey”) was developed to assess helmet use behavior among undergraduate students. Most of the questions were multiple-choice or used a Likert scale. Some questions had a write-in response option, and the final question was an open-text response that allowed participants to add general comments about the topics in the survey. Open-text responses were reviewed but not considered in analysis. Respondents were asked how often they wear helmets while riding their form of open-wheeled transport (OWT; such as a bike, scooter, or skateboard), as well as their reasons for wearing or not wearing a helmet. Information on their history of prior accidents involving OWTs and their form of OWT was sought, along with further questions to assess general knowledge of bike safety and risk perception of not wearing a helmet. Self-reported demographic details were also asked to ensure collection of a diverse and representative sample. To measure class years, participants were asked when they started their undergraduate career, and this was translated into how many years ago they began: 1 year represented freshmen at the time of the study, while 4+ years was mostly seniors. Freshmen (students in their first year) were compared to other students because the social culture at the university studied varies the most between freshmen and upperclassmen. Students were also asked where they had received information about helmet safety, to evaluate past helmet programs and identify avenues for promising pathways for reaching students.

#### Setting

Study data were collected using Qualtrics, a secure online survey platform. This survey was approved by the Stanford University Institutional Review Board (IRB), and the survey was closed early because the participant quota for compensation as outlined in the IRB-approved protocol had been reached (N = 400). The research team interviewed university healthcare providers, transportation and public safety staff, student research coordinators, and survey experts, who gave feedback on the survey design, recruitment strategy, and participant compensation.

#### Participants

Eligible participants were current Stanford University undergraduate students who used any form of OWT, both electric and non-electric, on campus. The study aimed to collect responses from a minimum sample size of 365 (derived from power calculations), and the research team secured funding to compensate 400 respondents. Information about the study and a link to the survey were sent to over 40 different student email lists and student dormitory group chats. Two incentives were offered: every respondent was entered into a raffle to win a free electric scooter and helmet; for every survey response, $2 was donated to the National Brain Tumor Society.

#### Analysis

Our hypotheses and statistical tests were specified prior to data collection. Additional *post hoc* tests conducted were exploratory, and results were intended to inform future research.

Our primary analyses were conducted using complete responses. After data collection, the research team consulted five students blinded to the data and asked them to group reasons for wearing or not wearing a helmet into four main categories: social factors, helmet characteristics, risk perception and knowledge, and other. The students were allowed to place each reason into multiple categories as deemed necessary.

It was hypothesized that students with electric OWTs wear helmets more frequently, because electric OWTs travel much faster than their non-electric counterparts. To evaluate this hypothesis, helmet usage frequency was compared between the two groups. Descriptive statistics for gender, race, year of starting undergraduate career, type of OWT, and whether or not the OWT was electric were reported as frequencies (percentage). Descriptive statistics were also used to summarize the distribution of frequency of helmet usage, reasons for wearing a helmet, and reasons for not wearing a helmet. When two categorical variables were compared, a Chi-Squared test was used, then Cramér’s V was used *post hoc* to further examine the association between the variables. When many variables were compared at once – for example, comparing the reasons for not wearing a helmet across different class years – statistical inference was based on 95% confidence intervals to avoid false positive results from multiple comparisons. When proportions in two different groups were compared – for example, the percentage of electric compared to non-electric OWT riders who never wear a helmet – a two-sample proportionality test was used. Data analysis was conducted using R version 4.3.1, and a p-value of <0.05 was considered to be statistically significant.

### Peer Agent Intervention

#### Participants

This intervention was approved by the Stanford University IRB, and informed consent was obtained from peer agents and students who pledged. Eligible participants for the peer agent were undergraduate students who always or nearly always wore helmets. We recruited 5 peer agents through email lists for student interest groups and dorms. As compensation, for each student they recruited, peer agents were awarded a $2 Amazon gift card (with a limit of $40 total) and they received a raffle entry for a $50 Bluetooth speaker. Of the recruited participants, 3 completed a 1-hour training session held by research staff. The training session described the motivation for the study, program logistics, and provided advice on how to engage students. The research staff led peer agents through example scenarios and provided feedback to the peer agents.

#### Pledge Intervention

After the training session, peer agents were instructed to talk to their friends, dormmates, and classmates about why they should wear a helmet. Each peer agent was asked to get at least 10 students to sign the pledge over a 5-week period. Students then completed a Qualtrics survey to document their pledge to wear a helmet and the name of the peer agent who talked to them. Every student who signed the pledge was compensated with a free helmet, bike light and a sticker. Three weeks after the intervention ended, a follow-up survey was distributed to students who pledged to ask about their helmet usage after signing the pledge.

#### Media Campaign

As the peer agents recruited students to sign the pledge, the research team conducted a 5-week general media campaign to promote helmet usage for all students. Based on the top reason for not wearing a helmet as reported in the survey, the research team created the slogan “Fashion Fades, Brain Injury Doesn’t: Helmet Up!” A research member created a campaign logo, which was a smiling brain fitted with a helmet. This logo, slogan, and facts about brain injury and helmets were featured in a flier posted across campus and distributed via email to student interest groups and dorms. Lastly, the research team wrote short messages (such as “Wear a Helmet!”, “Helmets = Brain Insurance”, or the campaign slogan) in chalk on sidewalks next to the student union.

#### Helmet Observation

To evaluate short-term changes in helmet use, members of the research team observed helmet usage in two areas of campus with high undergraduate commuter traffic and noted the numbers of cyclists, scooters, and skateboarders wearing or not wearing helmets. Researchers conducted 30-minute observation sessions twice daily, for a total of 6 sessions before and 6 sessions after the intervention. The number of students recorded was not fixed but instead depended on the rate of bicycle traffic, making this a Poisson sampling design. Using power calculations, it was determined that a minimum sample size of 311 was needed to detect a difference between the baseline and post-intervention observation periods (confidence level = 95%, margin of error = 5%). To avoid double-counting the same students, the observations across locations were conducted at different times.

#### Analysis

We compared the percentage of students seen wearing a helmet while riding an OWT at the baseline and after the intervention. A Kruskal-Wallis Chi-squared test was used, and a *p*-value <0.05 was considered statistically significant.

## Results

### Helmet Behavior Survey

A total of 400 completed student survey responses were recorded (**Table 1**), excluding 45 incomplete responses. More women (65%) were enrolled than men (31%), and the largest group of respondents identified as Asian (47%). This sample was generally representative of the Stanford population, but certain groups were over-represented: 52% of Stanford undergraduates were women, and 27% identified as Asian.^20^ All class years were represented, with the largest group (35%) being freshmen (the first year of a 4-year university system). The vast majority of participants used non-electric bikes (90%). There were few responses (N = 21, 5.25% of all respondents) to open-text responses, which is why they were thus excluded from analysis.

**Table 1.** Participant characteristics in the 2023 Caring for Your Brain Survey (N = 400)

|  | N | % |
| --- | --- | --- |
| **Gender\*** |  |  |
| Woman | 262 | 65.5 |
| Man | 125 | 31.3 |
| Non-binary | 9 | 2.3 |
| Genderqueer | 6 | 1.5 |
| Non-conforming or gender fluid | 4 | 1.0 |
| Prefer not to state | 6 | 1.5 |
| **Race\*** |  |  |
| Asian | 187 | 46.8 |
| White | 126 | 31.5 |
| Hispanic | 58 | 14.5 |
| Black | 52 | 13.0 |
| Middle Eastern or North African | 22 | 5.5 |
| American Indian or Alaska Native | 10 | 2.50 |
| Native Hawaiian or Pacific Islander | 6 | 1.50 |
| Prefer not to state | 9 | 2.25 |
| **Years since starting undergraduate career** |  |  |
| 7 | 1 | 0.25 |
| 6 | 4 | 1.00 |
| 5 | 24 | 6.00 |
| 4 | 84 | 21.00 |
| 3 | 53 | 13.25 |
| 2 | 95 | 23.75 |
| 1 | 139 | 34.75 |
| **Type of OWT** |  |  |
| Bike | 361 | 90.25 |
| Scooter | 26 | 6.50 |
| Skateboard | 13 | 3.25 |
| **Electric OWT** |  |  |
| No | 362 | 90.50 |
| Yes | 38 | 9.50 |

\*For categories marked with an asterisk, participants were allowed to select multiple categories, so the sum of the column percentages exceeds 100.

Most students reported never or rarely (73%) wearing a helmet while riding their OWT (**Figure 1**). The percentage of responses for each frequency category was compared between electric and non-electric OWT users, and there were no significant differences for any of the frequency categories (p=0.43 for daily, p=0.44 for a few times a week, p=0.77 for a few times a month, p=0.23 for rarely, p=0.15 for never; two-sample proportionality test) (**Figure 1**). When helmet usage was stratified by class year, there was no significant difference between the class years in their reported helmet usage frequency (p = 0.29, Chi-squared test) (**Figure 2**). Helmet usage and class year were weakly correlated (Cramér’s V = 0.24). The most common sources of information were parents (68%), medical providers (36%), and friends (33%), while the most common university source was New Student Orientation (25%), in which freshmen are introduced to the resources and culture on campus.

**Figure 1:**
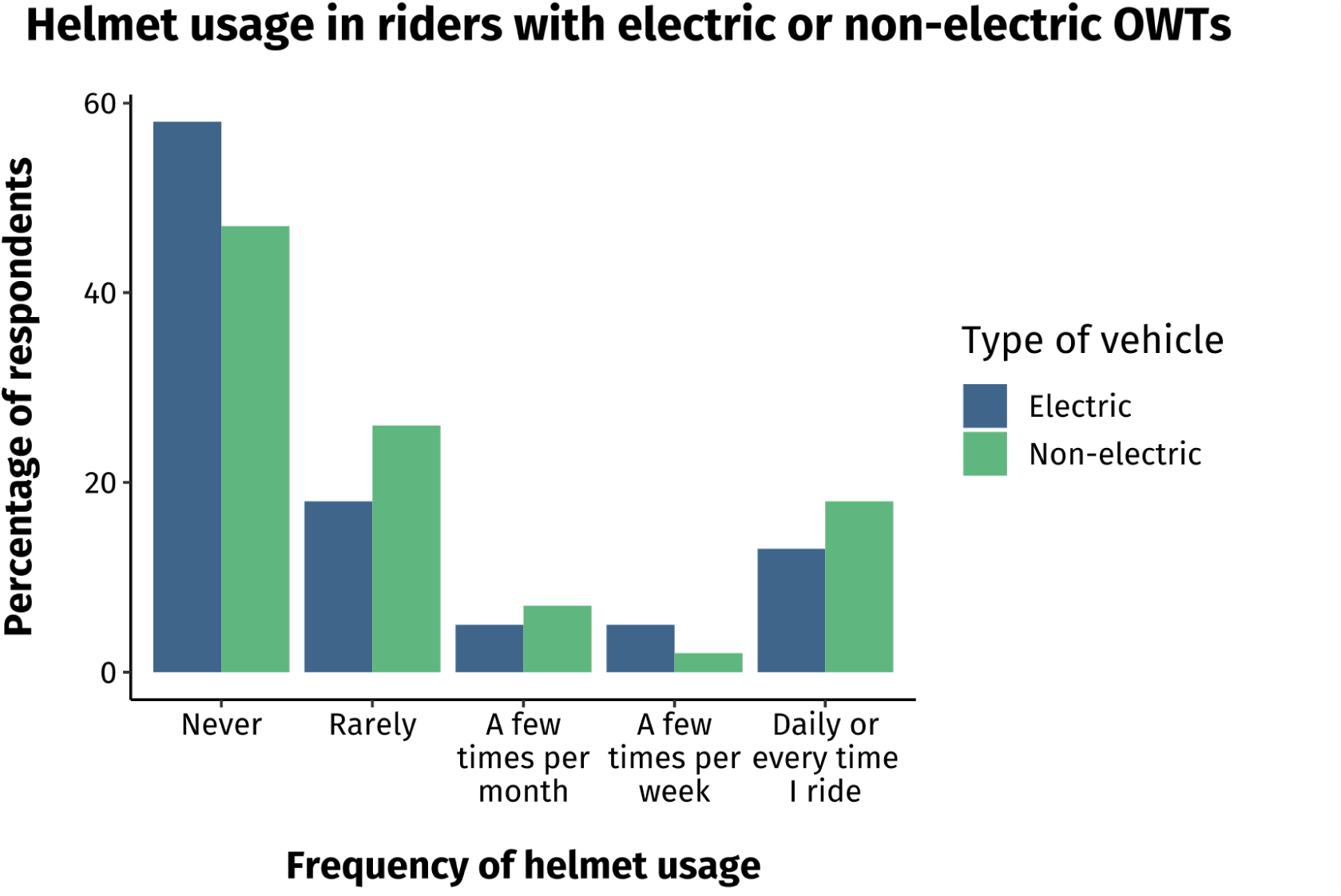
Responses to the question “How often do you wear a helmet while riding your OWT?” (N=400), stratified by users with electric and non-electric OWTs. OWT: Open-wheeled transportation.

**Figure 2.**
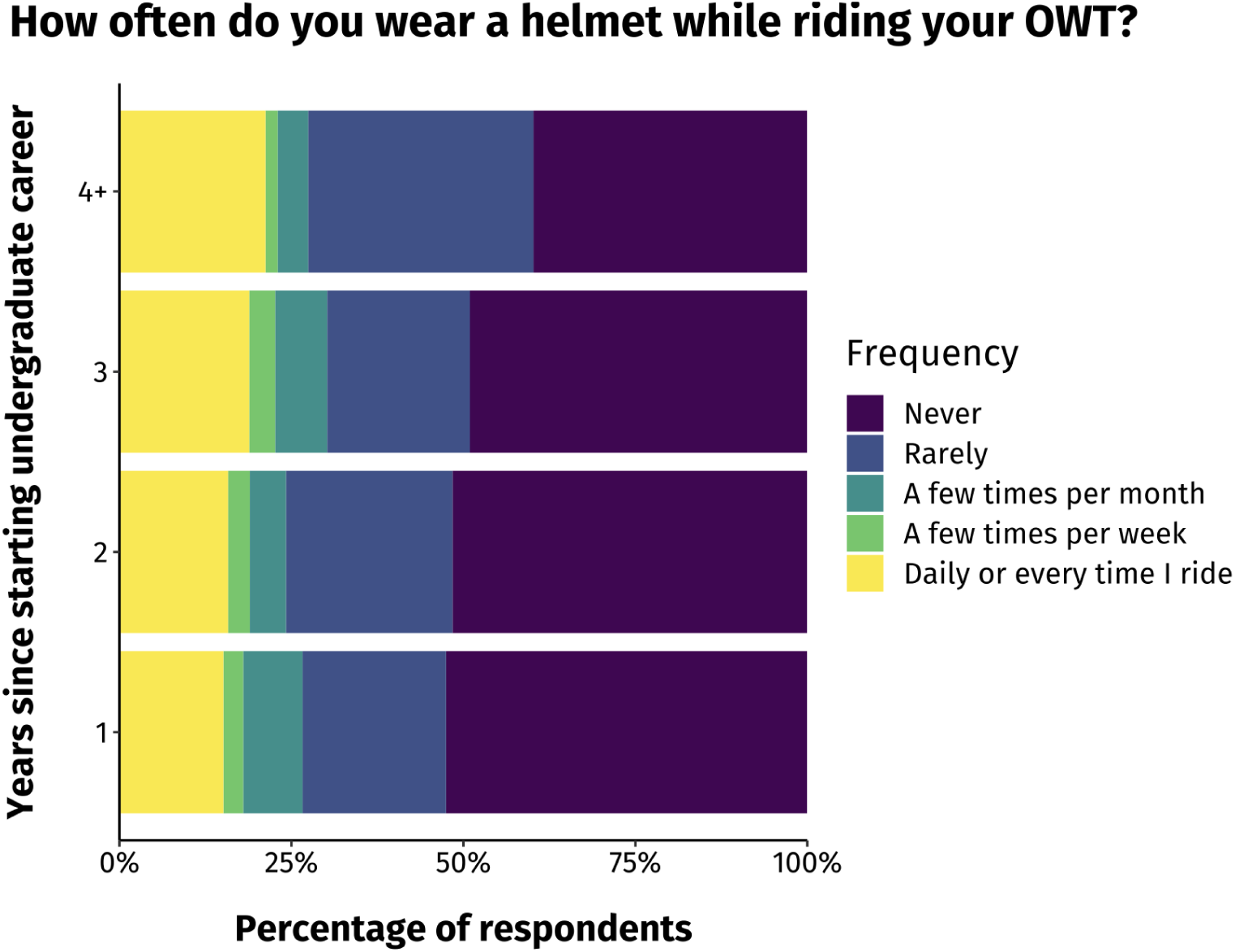
Responses to the question “How often do you wear a helmet while riding your OWT?”, separated by class year (N=400). OWT: Open-wheeled transportation.

The main reasons for not wearing a helmet involved characteristics of the helmet, social influence, and risk perception **(Figure 3)**. None of the reasons for not wearing helmets were homogeneously selected by the different class years **(Supplemental Figure 1)**. The most common reason for non-freshmen was “Helmets mess up my hair” (48%) while the most common reason for freshmen was “It’s unlikely I will fall/crash” (42%). The most common reasons for not wearing a helmet were sorted into risk perception categories (**Figure 4)**. Additionally, a higher percentage of students reported that they usually or often wear a helmet when riding off campus (55%) than riding a longer distance on campus (25%) or to class (21%).

**Figure 3.**
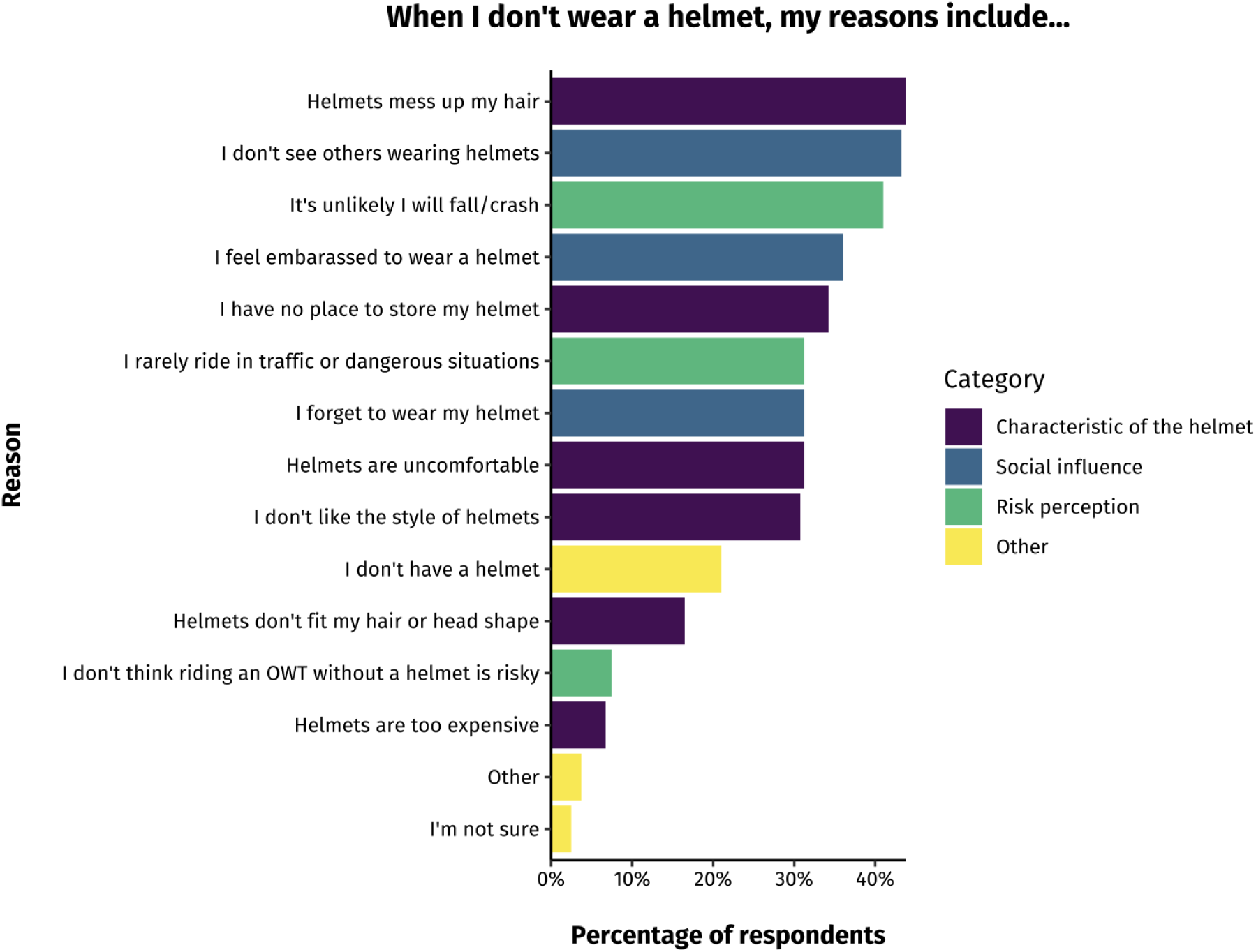
Responses to “When I don’t wear a helmet, my reasons include…”, separated by category (N=400). These responses were sorted into categories by several students blinded to the data. Two reasons were sorted into two categories. “I have no place to store my helmet” was sorted into “Characteristic of the helmet” and “Other,” while “I forget to wear my helmet” was sorted into “Social influence” and “Other.” In the figure, they were color-coded according to their dominant category, because “Other” is less informative for interpretations.

**Figure 4.**
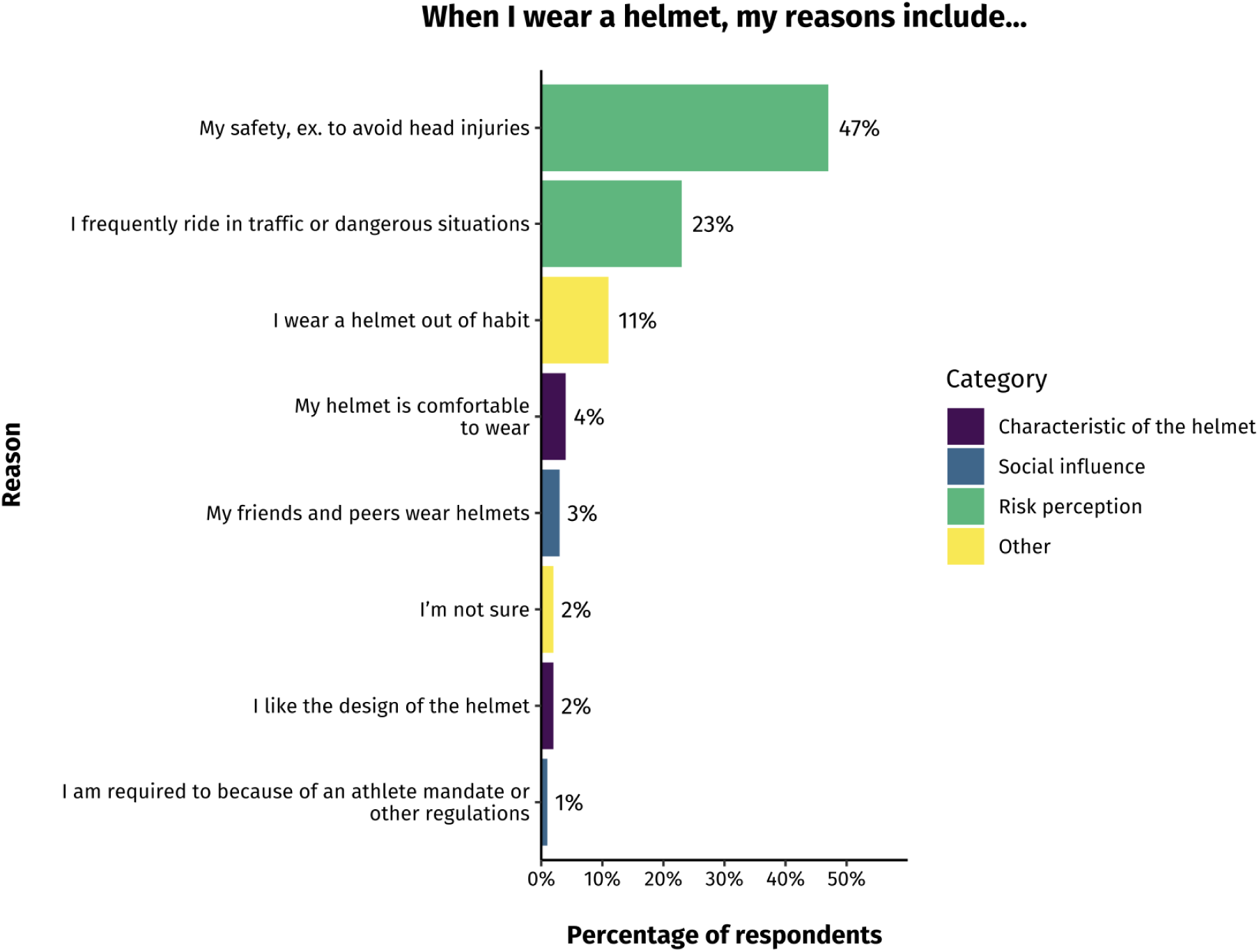
Responses to “When I wear a helmet, my reasons include…”, separated by category (N=400).

### Peer Agent Intervention

In the pilot intervention, 5 peer applications were received, and 3 peer agents completed the training. They recruited a total of 13 student pledges (although notably, 12 student pledges were from the same peer agent). The observers recorded a total of 4885 bike, scooter, and skateboard rides (N=2715 before the intervention, and N=2170 after the intervention). There was no significant difference in the percentage of helmet usage before and after the intervention (**Figure 5**, p = 0.48, Kruskal-Wallis Chi-squared test), however, there was an upward trend, potentially underpowered by the low number of pledges received in this pilot study. Of the 13 students who pledged, 6 responded to the 3-week follow-up survey. Before the intervention, 67% of respondents never or rarely wore a helmet, and after the intervention, 100% wore a helmet a few times a week or daily. 83% of respondents agreed or strongly agreed with the statement “Since signing the pledge, I changed my helmet usage because of the interactions I had with a peer agent.”

**Figure 5.**
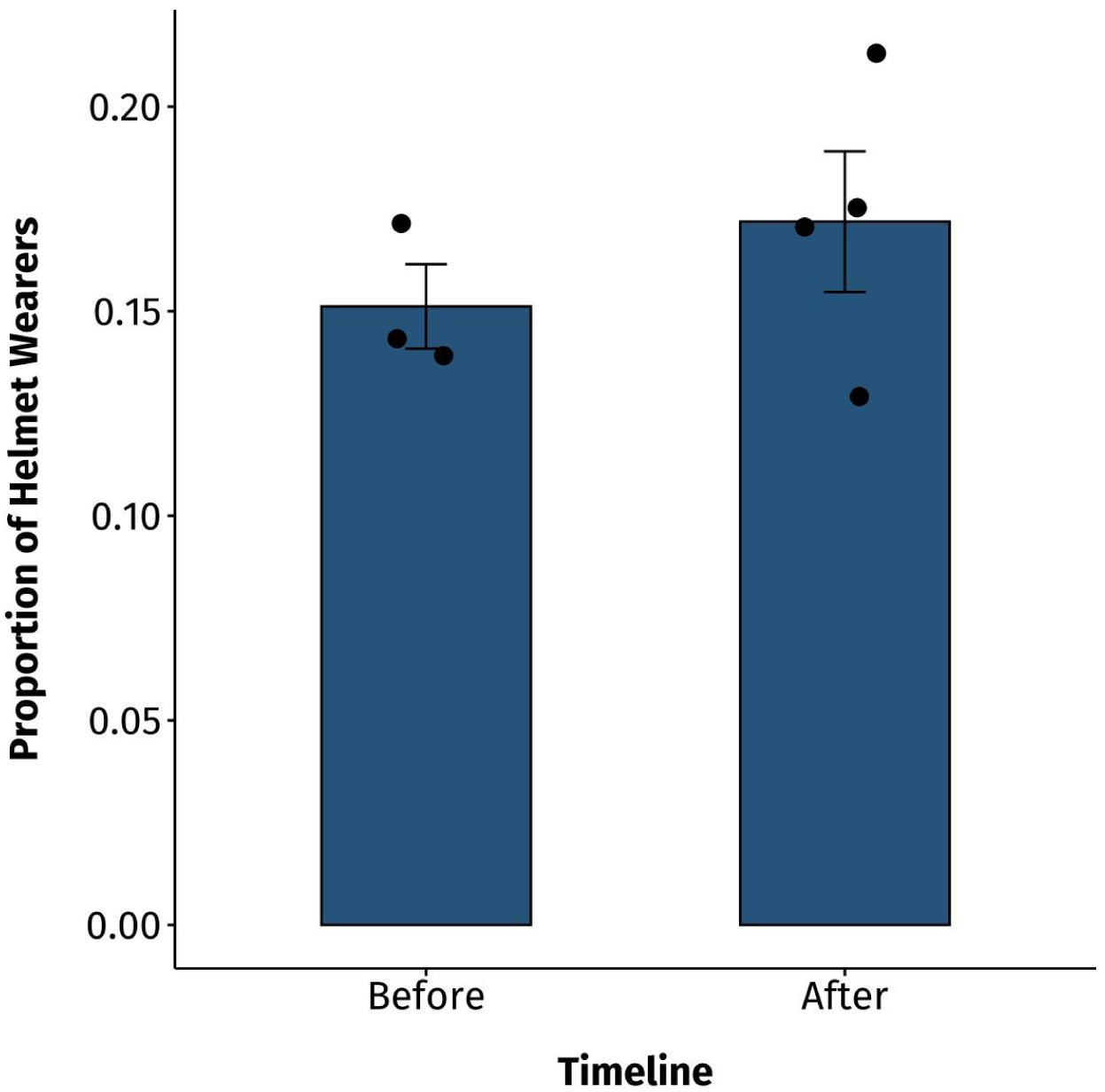
Proportion of helmet wearers before and after the 5-week pilot intervention. Data points represent the average proportion of helmet wearers per day of recording, and means across days are visualized with standard error bars.

## Discussion

This study surveyed undergraduate students with electric and non-electric OWTs about their helmet behavior and attitudes. We expected that students with electric OWTs would be more likely to wear helmets, because these vehicles often travel faster than non-electric forms of transport, and are more likely to result in concussions, severe brain injuries, and internal injuries.^3,4,21^ However, our findings are concerning because despite the greater relative risks of electric bicycles and scooters, we found no difference in helmet usage of electric and non-electric OWT users – both report alarmingly low frequencies of helmet usage. All class years reported a similarly low helmet usage, yet class years selected different reasons for why they do not wear helmets. Based on the reasons why students do not wear helmets – a mix of social influence, characteristics of the helmet, and risk perception – we found a multi-faceted intervention in the literature and aimed to validate it. We piloted an intervention and demonstrated a behavior change in the students who pledged to wear a helmet, but we did not document a larger change in helmet behavior across campus, likely because of the low number of peer agents recruited. Nevertheless, we observed an upward trend in helmet usage that reflects potential success in future interventions.

The survey replicated previous findings that the majority of undergraduates at the studied university report never wearing helmets.^8^ Both surveys found that students are less likely to wear a helmet on campus than off campus, and they perceive their risk of a crash to be low. This study’s finding that undergraduate students do not wear helmets because they mess up hair confirms previous research that the appearance of a helmet was a major reason why teenagers refused to wear one.^14^ However, some of the survey results differ from past findings. In the present study, the most common reasons for not wearing a helmet were “Helmets mess up my hair,” “I don’t see others wearing helmets,” and “It’s unlikely I will fall/crash.” In contrast, other US surveys found different reasons to be most prevalent, such as inconvenience, not owning a helmet, and forgetting to wear one.^8,12,16^ A previous survey from the same university did not find social influences to be among the top reasons.^8^ This could be because previous surveys did not include as many answer choices involving social influences, so participants did not have as many opportunities to report this reason. In general, this survey replicated previous trends in helmet behavior, indicating external validity.

The intervention resulted in an upward, although non-significant, trend in helmet usage: the students who pledged reported a change in behavior, indicating that the pledge was successful 3 weeks after the intervention. However, our intervention did not successfully replicate the original peer agent program because we did not observe a significant increase in helmet usage after the intervention, likely due to low recruitment of peer agents and students to pledge, resulting in low statistical power. Ludwig and colleagues recruited 15 bicyclists, and while we had aimed to recruit 20 students, we only received 5 applications. This could be because (as seen from the survey results) there is a small proportion of students who regularly wear helmets, even smaller than Ludwig and colleagues, who observed that 40% of students wore helmets before the intervention.^18^ Thus, there was a small pool of helmet-wearers to recruit from. Additionally, members of the research team had conversations with students who wore helmets regularly but did not want to become a peer agent. These students noted their unsuccessful prior attempts to persuade friends, suggesting that helmet advocates at universities may feel discouraged. Future interventions should provide additional support to help them address these challenges. Given our survey findings that helmet usage is a problem across class years and different class years reported different barriers to helmet usage, we recruited from more than one class year: our peer agents were first year and third year students. Further research should be conducted to determine if there are class year differences in the effectiveness of the peer agent program. We also found it helpful to offer free helmets to students who pledged, and recommend that future interventions incorporate this. The peer agent program is a targeted way to identify students who need a helmet and are committed to wearing one, thus increasing helmet accessibility and usage.

### Implications for Interventions

There was no difference in helmet usage between students with electric and non-electric open-wheeled transport (OWT), indicating that future interventions should address the lack of risk perception regarding electric scooters, bikes, and skateboards through the use of targeted safety messages. Future interventions should also cater to individual class years. Reasons for not wearing helmets were not homogeneously selected by the different class years, underlining the importance of designing interventions that target the needs of each class year. For instance, a higher proportion of younger class years (1-2 years since starting university) reported not wearing helmets due to risk perception reasons, which could be incorporated into future interventions targeting those class years.

This study also suggests potential avenues to reach students, and which messages should be emphasized. Parents were the main source of helmet safety information for students, though if university groups struggle to reach parents, they should also consider events during freshmen orientation. These events should endeavor to compel students to care more about their safety than their hair or social acceptance, because the students in this study who wore helmets did so because of their safety.

While social interventions and education may be helpful, researchers comparing the efficacy of legislative and educational efforts found that while both increased bike helmet wearing, legal mandates were most effective.^22^ Mandates may be particularly useful at universities, because younger groups are less risk-averse than older groups^23,24^ and past legislation (on the minimum drinking age, for example) successfully addressed age-related risk perception.^25^ Future researchers should evaluate how to overcome the barriers accompanying mandates, such as proposing policies to aid enforcement. For example, the state of Washington increased seat belt usage from 83% to 93% when the Chief of the Washington State Patrol made seat belts a law enforcement priority, and the state participated in a media campaign.^26^ Research on how to mitigate other obstacles is necessary to ensure successful implementation of helmet mandates.

### Limitations

The study has limitations, such as in the recruited sample. Many of the email lists used to recruit participants were pre-medical or science interest groups, which could skew results if pre-medical students are more risk-averse or knowledgeable about helmets than others. Additionally, the sample for the helmet attitudes and behavior survey may not have been representative of the Stanford undergraduate population, as women and Asian students were overrepresented. Due to challenges with recruitment, the pilot peer agent study was likely underpowered for detecting changes in the percentage of helmet usage after the intervention.

### Conclusion

This survey provides an in-depth look at the nuanced reasons why undergraduates do not wear helmets, and suggests initiatives to make universities safer. Undergraduate students reported not wearing helmets due to social influences, a lack of risk perception and knowledge, characteristics of the helmet, and other factors. Interventions must address multiple barriers experienced by students, such as counteracting negative peer pressure through the use of peer agents. However, mandates could also be instrumental to promoting helmet usage. Increasing helmet usage on college campuses will not only prevent debilitating injuries, but also promote the safety of the broader community.

## Transparency, Rigor, and Reproducibility Summary

This study was approved by the Stanford Institutional Review Board. This study was not pre-registered in a repository because it had been extensively reviewed by institutional committees. The study on surveying helmet usage aimed to collect responses from a minimum sample size of 365 (derived from power calculations with an alpha of 0.05 and a power of 0.80), and the actual sample size was 400. The study on examining the effectiveness of the peer agent program aimed to observe 311 bikers (derived from power calculations with an alpha of 0.05 and a power of 0.80), and the actual sample size was 4885. There were 5 students who applied to the peer agents program, and 3 who completed the training. They recruited a total of 13 students, 6 of whom responded to the follow-up survey. Students assisting the research team were blinded to the data when they grouped reasons for wearing or not wearing a helmet into hypothesis categories. Our hypotheses and statistical tests were specified prior to data collection. Data analysis was conducted using R version 4.3.1, and a *p*-value of <0.05 was assigned to statistical significance. When many variables were compared at once, statistical inference was based on 95% confidence intervals to avoid false positive results from multiple comparisons. The survey questions are available in the supplementary materials.

## Supporting information

Supplement_Survey Questions

## Data Availability

All data produced in the present study are available upon reasonable request to the authors.

## Acknowledgements

We thank the Synapse Undergraduate Research Group (SURG) members for their contribution to the literature review, study design, and participant recruitment. We thank Ariadne Scott and our other collaborators from the Stanford Transportation and Public Safety Departments for their feedback on our study design and preliminary results, and for giving access to their relevant data. We thank the Camarillo lab members for their feedback on preliminary results. We also thank the other stakeholders interviewed – healthcare providers, campus transportation and public safety staff, student research coordinators, and survey experts – for their suggestions on improving our study design. We also thank the participants for contributing their time to the study.

## Footnotes

### Contributors

AD and GL developed the research idea, and AD, GL, ELF, and DC contributed to the study design. AD, GL, and SC conducted data analysis under the guidance of VR. AD, GL, and SC designed the figures, which were edited by VR and DC. AD, GL, and SC wrote the manuscript, which was edited by ELF, VR, and DC. All authors have read and approved the final manuscript.

