## Supplement_Survey Questions for "Promoting helmet usage in undergraduates with electric and non-electric bicycles and scooters: a single-site survey and intervention"

1. Have you ever (before or during your college career) been in an accident in your OWT?
    - Yes
    - No
2. What was the severity of your accident? If you have been in multiple accidents, please only consider the most severe accident.
    - Mild (no or very minor injuries)
    - Moderate (bruises, swelling, or minor scrapes)
    - Severe (broken bones, dislocated limbs, skull injury or concussion, or unconsciousness)
    - Prefer not to answer
3. Which OWT do you most frequently use on campus? (select one)
    - Bike
    - Skateboard
    - Scooter
    - Rollerblade
    - Other (short answer)

*The following questions are based on your primary OWT, which was selected in the previous question.*

4. Is your OWT electric?
    - Yes
    - No
    - I don't know
5. How often do you wear a helmet while riding your OWT?
    - Never
    - Rarely
    - A few times per month
    - A few times per week
    - Daily or every time I ride
6. When I wear a helmet, my reasons include … (select all that apply)
    1. I frequently ride in traffic or dangerous situations
    2. My safety, ex. to avoid head injuries
    3. I am required to because of an athlete mandate or other regulations
    4. My friends and peers wear helmets
    5. I like the design of the helmet
    6. My helmet is comfortable to wear
    7. I wear a helmet out of habit
    8. I'm not sure
    9. Not applicable (I don't own a helmet, or never use it)
    10. Other: (short answer)
7. When I don't wear a helmet, my reasons include … (select all that apply)
    1. I rarely ride in traffic or dangerous situations
    2. It's unlikely I will fall/crash
    3. I don't think riding an OWT without a helmet is risky
    4. I don't see others wearing helmets

    5. I feel embarrassed to wear a helmet
    6. I don't have a helmet
    7. I have no place to store my helmet
    8. I forget to wear my helmet
    9. I don't like the style of helmets
    10. Helmets are too expensive
    11. Helmets mess up my hair
    12. Helmets are uncomfortable
    13. Helmets don't fit my natural hair, braids, hair accessories, or head shape
    14. I'm not sure
    15. Other (short answer)
8. I usually/often wear a helmet in the following occasions: (select all that apply)
    - Riding off-campus, such as in traffic or on bike paths
    - Riding to class
    - Riding a short distance on campus, such as to a dorm or dining hall
    - Riding for a longer distance on campus
    - None of the above
    - Other (short answer)
9. I think wearing a helmet is …
    1. Not at all important
    2. Somewhat important
    3. Important
    4. Fairly important
    5. Very important
    6. No opinion
10. How concerned are you about acquiring a brain injury due to not wearing a helmet while riding an OWT?
    1. Not at all concerned
    2. Somewhat concerned
    3. Concerned
    4. Fairly concerned
    5. Very concerned
    6. No opinion
11. How well do you understand the risks of not wearing a helmet while riding?
    - I don't know why I should wear a helmet
    - I know some of the risks of not wearing a helmet
    - I know a lot about the risks of not wearing a helmet
12. I have gotten information about helmet safety from … (select all that apply)
    - Parents
    - Friends
    - Medical professionals, inside or outside of Stanford
    - Vaden Health Care
    - Dorm staff
    - Approaching Stanford
    - NSO (New Student Orientation)
    - Stanford Athletic coach, Stanford Athletics

- Stanford Public Safety (ex. bike safety classes after citations)
- Stanford Transportation, Bike Program (ex. dorm roadshows)
- Stanford Daily
- Other (short answer)
- None of the above

13. Are you a Stanford varsity athlete (not club or intramural)?
    - Yes
    - No
14. [If yes to being a varsity athlete]
    - Which varsity sport do you play?
        - Drop down menu of all 36 sports
    - Is your team required to wear a helmet outside of sports (when on a bike or open-wheeled transport)?
        - Yes
        - No
        - I don't know
    - [If yes] On a 5-point scale from 1 = strongly disagree and 5 = strongly agree, how much do you agree with the following statements?
        - I follow the helmet mandate.
        - I support the mandate and think it is beneficial.
        - I think the helmet mandate is helpful in making athletes safer.
15. What is your email?
16. What year did you start your undergraduate career at Stanford?
    - 2016
    - 2017
    - 2018
    - 2019
    - 2020
    - 2021
    - 2022
    - Other:
17. I most often describe my gender as … (We ask this to make sure we recruit a diverse and representative population.) Select all that apply.
    - Woman or woman-identified
    - Man or man-identified
    - Non-Binary
    - Genderqueer
    - Gender variant/Non-conforming/Gender fluid
    - Other : ____________________________________________
    - Prefer not to state
18. I most often describe my race as … (We ask this to make sure we recruit a diverse and representative population.) Select all that apply.
    - Black or African American (e.g. African American, Jamaican, Ethiopian, Haitian, etc.)
    - American Indian or Alaska Native (e.g. Navajo Nation, Blackfeet Tribe, Mayan, Aztec, etc.)

- Asian (e.g. Chinese, Vietnamese, Indian, etc.)
- Native Hawaiian or Pacific Islander (e.g. Kanaka Maoli, Samoan, Chamorro, etc.)
- Middle Eastern or North African (e.g., Lebanese, Iranian, Egyptian, Moroccan, Israeli, Palestinian, etc.)
- Hispanic, Latinx, or Spanish Origin (e.g. Puerto Rican, Cuban, Salvadoran, etc.)
- White (e.g. German, Irish, Italian, etc.)
- Other: ______________________________________________
- Prefer Not to State

19. Please use this space to share any additional comments about the topics of this survey (ex. attitudes about helmets, brain injury, campus culture). (Optional)
