## Supplemental Figure 1 for "Promoting helmet usage in undergraduates with electric and non-electric bicycles and scooters: a single-site survey and intervention"

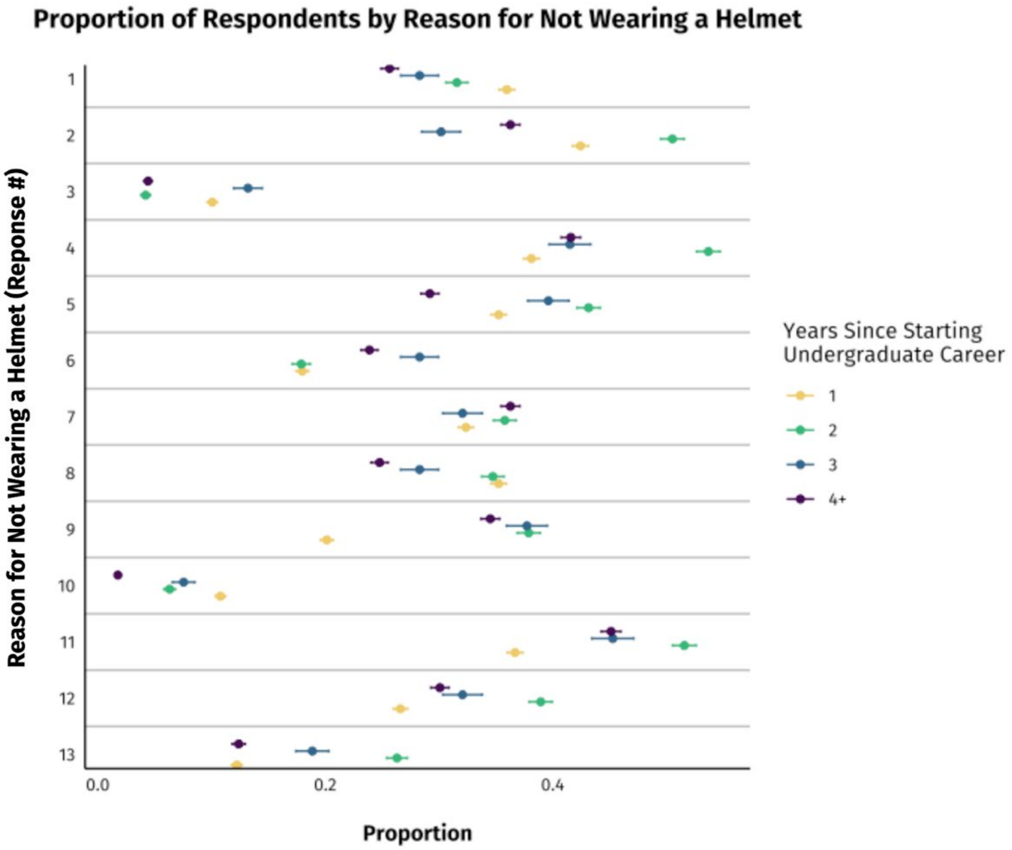


**Supplemental Figure 1.** Proportion of respondents that selected each reason for not wearing a helmet, separated by class year (i.e. years since starting undergraduate career) (N=400). The reasons for not wearing a helmet are numbered on the y-axis, and the key is located in the appendix (Survey Questions, Question #7).
